# Discriminative touch for manual action in chemotherapy induced peripheral neuropathy

**DOI:** 10.1101/2025.03.05.25323401

**Authors:** RD Roberts, W Chua, A Khatibi, NP Holmes, C Palles, R Kussaibati, AM Wing

## Abstract

Chemotherapy induced peripheral neuropathy (CIPN) is a frequent side effect of a number of chemotherapeutic agents which are widely used in the treatment of common cancers. Sensory symptoms primarily affect the fingers and toes and include numbness, tingling and pain. However, there is limited understanding of how CIPN may impact manipulation skills. Questionnaire and focus group methods were used to explore the experience of CIPN side effects impacting manual activities in 25 self-selected participants recruited from a cancer charity web site advertisement. Participants’ responses demonstrated varying degrees of impact of touch impairment associated with CIPN on the performance of manual activities involving bimanual, unimanual and directed touch. Examination of the components of the affected activities, together with participants’ reports of alterations in their experience of touch, was used to generate hypotheses about why some manual tasks are more affected than others. In future research it is proposed that quantitative measures of manual activities should complement patient reported outcome measures in evaluating the mechanisms underlying issues in sensory motor control of manual activities caused by the effects of CIPN.

## Introduction

Chemotherapy induced peripheral neuropathy (CIPN) is a common side effect of a number of chemotherapeutic agents including taxanes and platinum analogues, both of which are widely used in the treatment of common cancers such as breast, prostate, lung and gastrointestinal malignancies. CIPN affects upwards of 60% of patients with the effects being more pronounced with platinum analogues (Burgess et al., 2021). Sensory symptoms primarily affect fingers and toes (stocking and glove syndrome) and include numbness, tingling (paraesthesia), unpleasant sensation (dysesthesia) and pain following stimuli that do not normally cause pain (allodynia). These symptoms tend to persist after termination of chemotherapy even, in some cases, worsening for a period of time (coasting), and can severely compromise patients’ quality of life. Different physiological mechanisms for CIPN are proposed to underlie various chemotherapy agents, but, simply stated, in each case afferents between tactile mechanoreceptors and the spinal cord are involved through changes in synaptic or nerve conduction properties (Burgess et al., 2021). The occurrence of CIPN may lead to dose reductions, treatment alterations and early cessation of chemotherapy which can ultimately lead to poorer outcomes.

Detection of the onset of CIPN often draws on National Cancer Institute Common Terminology Criteria for Adverse Events (NCI-CTCAE). However, it has been suggested that this is not a reliable tool for clinical research due to significant inter-rater variability and a lack of sensitivity to changes in CIPN over time (Cavaletti et al., 2010; Colvin, 2019). For clinical research into the progression of CIPN symptoms, questionnaires with patient reported outcome measures (PROMs) have been widely used. Li et al. (2022) reviewed CIPN questionnaires utilising PROMs on a number of dimensions including responsiveness, validity and reliability. For example, the European Organization for Research and Treatment of Cancer (EORTC) CIPN20 questionnaire (Postma et al., 2005) was highly rated for responsiveness to CIPN symptom development. It also scored well for detailed “proximal” outcomes (direct effects on actions such as walking problems, due to difficulty feeling the ground underfoot). In the present study we use PROMs, supplemented by focus groups, to examine the reported experience of CIPN, what manual tasks are affected and how participants coped with the impact of CIPN. We aimed to map these reports onto current understanding of sensorimotor control based on underlying biomechanical, neurophysiological and psychological mechanisms. In the longer term, our approach may contribute to asking more detailed questions about manipulation skills. Such an approach aims to facilitate the development of common principles to explain a seemingly diverse set of tasks impacted by CIPN. We also ask whether there might be a place for objective tests that bridge the gap between elemental passive touch tasks, such as pressure and vibration detection, and complex activities such as writing and buttoning skills.

In order to elaborate on sensorimotor control mechanisms, consider writing (see Figure 1). Taking hold of a pencil in a tripod grip comprising thumb, index and middle fingers activates pressure sensitive mechanoreceptors in the smooth (glabrous) skin of the pads of each digit. Neural signals from these sensors are relayed by afferent fibres to the spine and then to the brain where a cortical representation of the pencil’s location in the hand is formed. Writing with the pencil involves pressing the pencil on the paper. Pressing down can cause the pencil to slip and change its location in the tripod grip. Even a small change in pencil position stimulates pressure sensitive, slowly adapting (SA1, SA2), mechanoreceptors. In addition, microslips against the pencil generate small vibration waves in the skin, activating rapidly adapting (RA1, RA2) mechanoreceptors. These afferent signals elicit supraspinal reflexes involving sensory and motor cortices and result in grip force adjustments to stabilise the pencil in the hand. Supraspinal reflexes are fast (of the order of 100 ms) but this delay could allow enough slip to make writing difficult or even result in dropping the pencil. An alternative to relying on supraspinal reflexes is to anticipate the effect of pressing down with the pencil and to increase grip force before any slip occurs. Writing includes a series of pencil strokes and each contact with, and movement over, the paper generates a series of tactile signals. In skilled manipulation, touch feedback from movement events is continually monitored against what would be expected so that consequences of inaccurate anticipation of forces or unexpected external disturbances can be quickly corrected (Johansson & Flanagan, 2009).

**Figure 1:**
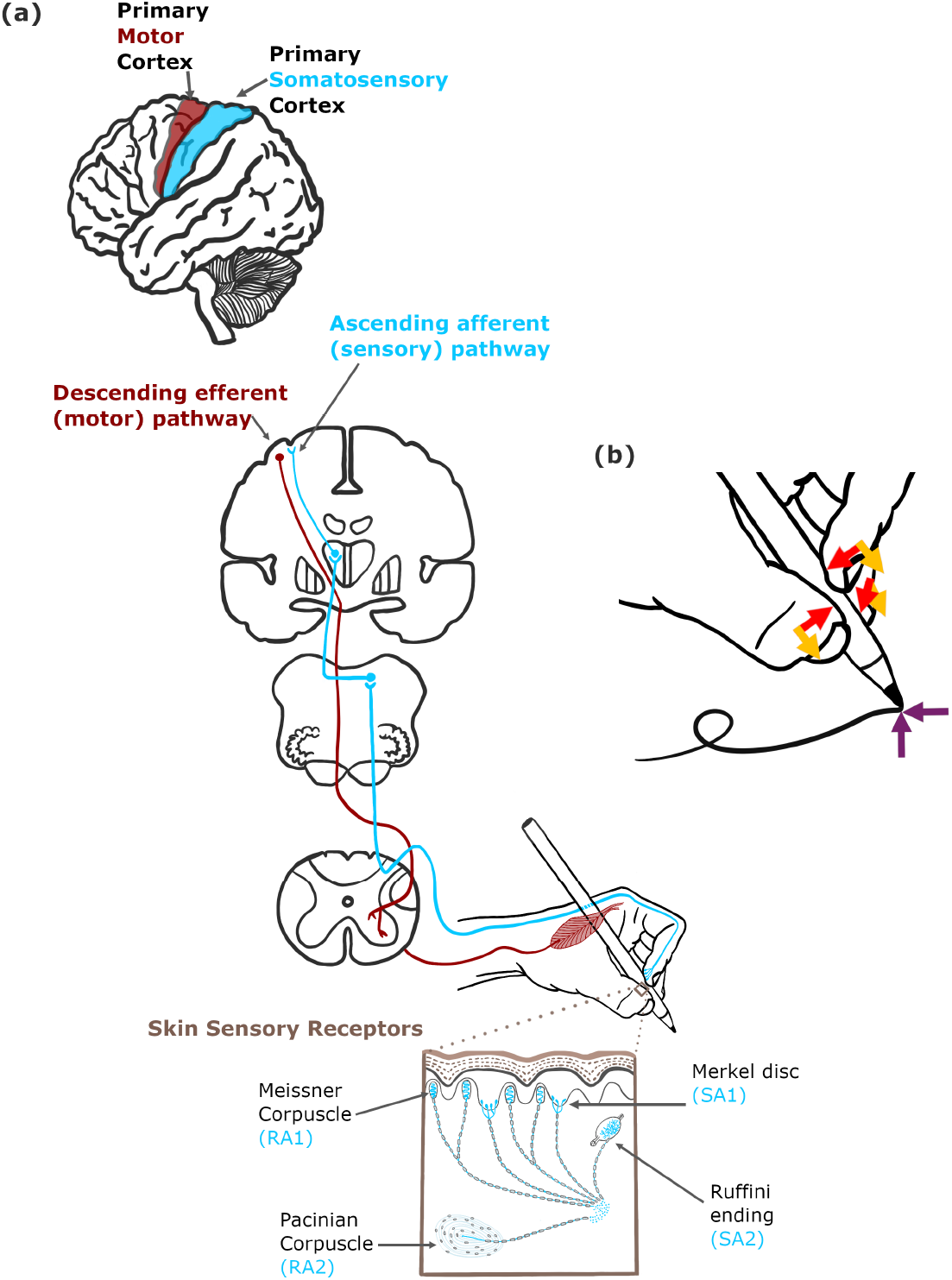
(a) Schematic of the ascending afferent (touch) and descending efferent (motor) pathways to and from the somatosensory and motor cortices in the sensorimotor control of writing grip. (b) In writing, the thumb, middle and index finger form a tripod. Grip force normal (shown by red arrows) to the surface develops friction between the pen and digits allowing tangential force (yellow arrows) to press the pen down on the paper without the digits slipping. Reactive forces to the pen tip, normal and tangential to the paper surface, are shown as purple arrows. Imbalance of the grip forces allows the pen tip to be pushed across the paper.

**Figure 2:**
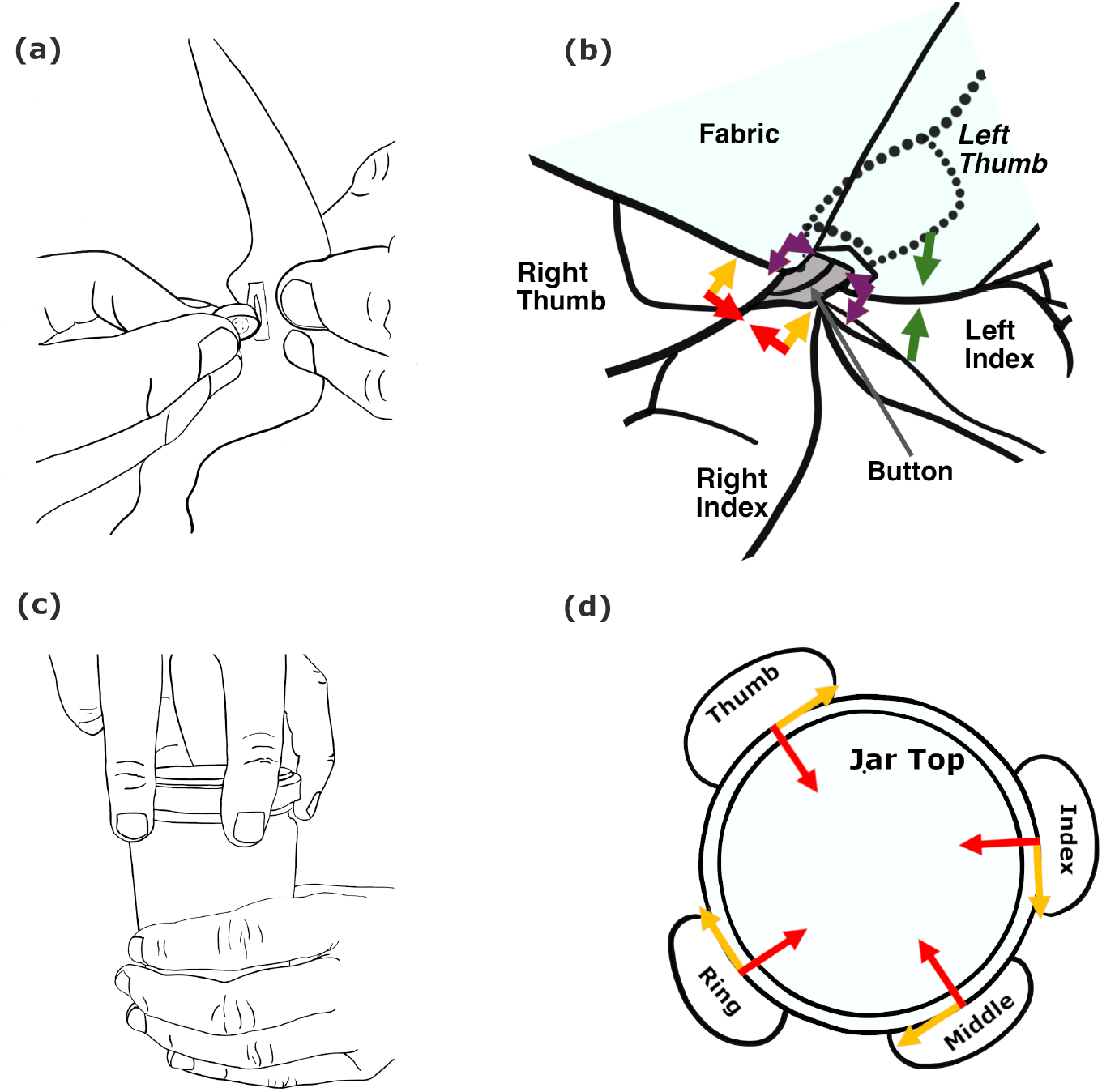
Grip and load forces in manipulation tasks: a) Buttoning a shirt. (b) The top-down view shows the right-hand thumb and index grip force normal to the button surfaces (shown by red arrows) develop friction allowing tangential force (yellow arrows) to push the button into the buttonhole without the digits slipping. Purple arrows here show reactive forces from the buttonhole) which has been opened using a pinch grip (normal forces shown by green arrows) pull of the material with the thumb and index of the left hand. (c) Opening a jar. (d) The thumb, index, middle and ring finger grasp and attempt to rotate the lid on the jar held by the other hand. Each digit exerts a normal force into the centre of the lid (shown by red arrows) which generates a friction to resist slipping due to the turning force (shown by yellow arrows) at each digit tangential to the lid. Tactile feedback from the multiple contact points is key to controlling grip and load forces needed to achieve these tasks.

## Methods

The study comprised two parts. The first involved questionnaires completed by a self-selected sample of people with lived experience of CIPN. The second part involved two focus groups drawn from a subset of those who completed the questionnaires. Ethical approval for the research was obtained from the University of Birmingham Science, Technology, Engineering and Mathematics Ethical Review Committee.

### Questionnaires

The participants were 25 adults (including author AMW) with lived experience of CIPN. Their average age was 62.7 (SD=8.0) years and 17 were female. They were recruited through an advertisement on the Cancer Research UK website.

Participants completed an online version of 3 questionnaires, which they accessed in Spring 2023. The first comprised the European Organization for Research and Treatment of Cancer Quality of Life Questionnaire (EORTC CIPN20, abbreviated here to QLQ; Postma et al., 2005). QLQ consists of separate questions for hand and foot relating to CIPN symptoms and activities experienced in the previous 7 days. The second questionnaire was the Patient Neurotoxicity Questionnaire (PNQ; Hausheer et al., 2006). PNQ has questions covering the previous 24 hours in terms of symptoms across upper and lower limb and a checklist of activities affected. The third questionnaire, which was developed for the study, is referred to as the University of Birmingham Questionnaire (UBQ). The UBQ questions are more focused on numbness and tingling than QLQ and PNQ but more open-ended in seeking responses to activities affected, coping strategies and extending the relevant timescale for reporting of symptoms back to the onset of CIPN. Together, these questionnaires encompass participants’ CIPN experience from its onset, which may have been during chemotherapy, up to the time of completing the questionnaires. Questionnaire details, including distribution of responses, can be found in the supplementary materials.

### Focus groups

Participants who indicated their interest while completing UBQ were invited to take part in an on-line focus group. Two focus groups of five participants were held in separate video conference (Zoom) sessions (A, B), each led by two of the authors (RDR, AMW) acting as facilitators and lasting one hour. The session began with a brief review of the questionnaire that the participants had completed 3 months previously, and a short presentation on neural pathways for touch (based on Witney et al., 2004). The facilitators then led off discussion of CIPN effects on manual activities by asking participants to write their name in pencil on a piece of paper and fasten a button on a shirt, items they had been asked to bring to the session. Participants were then invited to describe problems experienced in these and other manual activities and discuss strategies invoked to compensate for the impact of CIPN. The session concluded with discussion of sensory experiences of CIPN on the feet and how information reviewed in the focus group might be useful for others experiencing CIPN.

## Results

In our presentation of the results, we first summarise participant demographics and then report CIPN effects (in terms of both questionnaire responses and focus group statements) on hands and feet sensation, on writing and buttoning activities, and on other manual activities. In some instances, the questionnaire responses of a subset of participants are presented in terms of averages over responses to individual questions and across grades of response. A summary in terms of grading of questionnaire responses across all 25 participants can be found in the supplementary materials.

The focus group reporting is shown in tables with the sequence of columns indicating focus group (A or B), participant study ID, the time stamp (minutes, seconds) in the recording and participants statements. The statements are reproduced verbatim except for omissions of asides (…) and italicised parenthetical comments inserted by the authors. There is no rephrasing, and the original ordering is preserved as it represents raw data. It is grouped by themes agreed by the authors and confirmed with the participants. This conforms to standards suggested by Anderson (2010).

### Demographics

Based on the 25 participants’ responses to UBQ item 3, 68% (N=17) experienced numbness and tingling affecting the hands and the feet, 24% (N=6) just the hands and 8% (N=2) just the feet. The last two participants were excluded from the following analysis of the questionnaires as it focuses on the hand and the performance of manual activities. The demographics for the remaining 23 participants are shown in Table 1. Chemotherapy information was available for 15 of the 23 participants. Of these 10 received taxanes, 8 platinum compounds and 5 other agents, including Bortezomib, Lenvatinib, Pertuzumab, and Cyclophosphamide.

**Table 1.** Details of participants indicating numbness and tingling due to CIPN affecting the hands (N=23).

| **Measure** | **Average (SD)/Number** |
| --- | --- |
| Age (SD) | 62.5 (8.2) years |
| Female | 15 |
| Right handed | 20 |
| CIPN onset during chemotherapy | 17 |
| Years since chemotherapy (SD) | 4.3 (3.1) years |
| Number with CIPN improving | 5 |

### CIPN effects on hands and feet sensation

Averaging across QLQ items 1-18, 42% of responders experienced problems graded as 2 (“a little”), 3 (“quite a bit”) or 4 (“very much”). QLQ items 1-8 span symptoms (numbness, tingling, pain, cramps) in the hands separately from the feet. Items 9-15 (excluding item 10 which is ambiguous as to which body part it relates) cover six activities, three involving the hands and three the feet. Table 2 summarises the average percentage of respondents who graded these hands and feet symptoms and activities as 2 or above. PNQ items about symptoms do not distinguish hands and feet and so are not reported here. Although Table 2 shows symptoms involving the hand and feet were reported by equal proportions of the participants (50%), hand activities can be seen to have been more commonly affected (60%) than were activities involving the feet (20%).

**Table 2.**
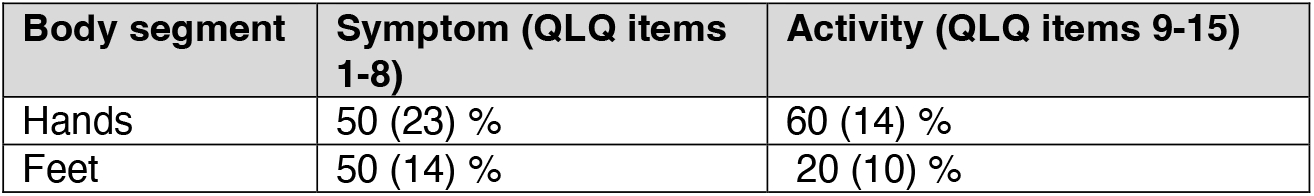
Mean (SD) percentage of respondents (N=23) assigning grades to QLQ items 1-8 (symptoms), 9-15 (activities) of the impact of CIPN at 2 or above grouped by symptom and activity for hand vs foot.

Responses to UBQ item 3 on numbness and tingling affecting hands and feet indicated 100% of participants experienced symptoms affecting the hands and 65% (N=15) had feet affected. This finding of higher percentages affected compared with those reported in Table 2 can be attributed to differences in the time periods covered by UBQ and QLQ. Specifically, QLQ refers to the week preceding completion of the questionnaires while UBQ covers the full period since chemotherapy. In response to UBQ item 9 about additional symptoms, 44% (N=10) of participants noted the nails were affected, while two participants (9%) commented on dry skin, and two on skin rash or deterioration.

Table 3 reproduces statements from the two focus groups elaborating on the sensory experience of the hand. The statements relate to numbness, tingling and to pain. Comments under numbness and tingling include reductions in CIPN symptoms. Under the pain heading, statements include reference to pain interfering with tasks in which fingertip pressure is required.

**Table 3.** Focus group statements relating to sensory experience: hand numbness and tingling and pain.

| Group | ID | Time | Statement |
| --- | --- | --- | --- |
|  |  |  | **Numbness, Tingling** |
| A | 25 | 27:55 | So I now, I really only have any loss of feeling right in my fingertips right at the very end of my fingers. So my feeling has improved. |
| B | 26 | 14:18 | Sometimes in the mornings I wake up, and I do this (*wiggles fingers*) to get them. I feel as if I've slept on them, even though I haven't slept on them, you know, because wake up with the pain and tingling, and I can't feel anything. |
| B | 11 | 16:30 | I had the tingle in my hands for a good 6 months after the chemo and they were actually trying to work out ... a better way to give me the chemo, so that I would have less problems with my hands ... because I use my hands to see with (*participant 11 is blind*). In the end they stayed with the normal once every 3 weeks, but once, once the tingling ... started, it just stayed. |
|  |  |  | **Pain** |
| A | 19 | 34:03 | There's an interference, isn't it? I think we're saying that there's some pain you can discriminate well enough. But now there's a pain element which is when you press. |
| A | 25 | 39:06 | Even now (*5 years on*), because the room is quite cool my fingertips are painful and so to take anything I know I'm touching because of the pain, more so than the sensation of touching. |
| A | 19 | 56:54 | I'm wearing gloves in the morning, and also for the discomfort If I was to touch something even just hooking the dogs lead on and off, trying to use the clasp and things like that can be uncomfortable. |
| B | 10 | 03:59 | I've got ... few problems with my hands ... the only thing I can't do is play the guitar because of the pressure on the very tips of my fingers, and I just can't do it now. ... yes, it's pain. |
| B | 11 | 04:19 | I find it painful to read very new Braille for a long time because when it's ... it's sharper. and so ... I could only read it for so long, and then it would get painful, whereas if it was older, Braille is duller ... it was easier to read. |

Focus group statements about sensory experience of the feet are shown in Table 4. As in Table 3 the statements are grouped under numbness and tingling and, separately, for pain.

**Table 4.** Focus group statements relating to sensory experience: foot numbness and tingling and pain.

| Group | ID | Time | Statement |
| --- | --- | --- | --- |
|  |  |  | **Numbness and tingling** |
| A | 9 | 52:47 | My feet are much more sensitive to what's under the sole. in fairly cold water ... initially it felt as if I were walking through seaweed ... mixed sensations around my ankles, which eventually I adapted to and got used to. |
| A | 9 | 53:57 | ... it's much more the level of the feet, and you know, and I have tingling when I'm lying in bed. |
| B | 11 | 25:46 | I've had numbness in my toes .. ever since I had chemo (*6 years ago*) ... occasionally I'm aware of balance not being right. But it's ... manageable. But it's been there for well, for 6 years. |
| B | 10 | 26:37 | trip quite a lot at the beginning. and I did fall down 3 stairs as well in in the house once, and I've taken avoiding strategies for that. Really, I walk more slowly, and that has solved the tripping problem. I never wear shoes with any sort of heel on them, because it was a heel I caught coming down the stairs, and I always hold a rail, either in the house coming down the stairs or when I'm out. |
|  |  |  | **Pain** |
| A | 25 | 51:31 | It's the pain for me in the cold. ... I've got to have my foot on the clutch for any length of time. ... The pressure brings on too much pain to concentrate on driving. So at least with an automatic, you can have one foot free. |
| A | 25 | 52:19 | So I never stand still either. So if I'm in a queue for something, I don't stand still. I can't, because the pain is too great. |
| B | 24 | 23:24 | My left foot ... is affected and has been ever since I started chemotherapy (*3 years ago*)... a burning sensation .. when stepping up, down stairs or steps. I'm very conscious, as I can't always feel the ground properly. |
| B | 10 | 28:49 | I wear bed socks, thick bed socks every night in bed because I've got allodynia, which is this sort of intensive sensitivity of my feet and that helps. |

### Writing and buttoning activities affected by CIPN

Table 2 showed a majority of participants reported in QLQ that one or more of three hand activities were affected by CIPN. The three activities which caused difficulty are listed, with associated percentages of participants affected, in Table 5. They were (i) holding a pen, which made writing difficult, (ii) manipulating small objects with the fingers, for example, fastening small buttons, and (iii) opening a jar or bottle. The distribution of gradings for these three activities in QLQ across all 25 participants may be seen in Table (S2) in the supplementary materials.

**Table 5.** Responses to QLQ indicating percentage reporting activity affected.

| Activity | Percent affected |
| --- | --- |
| Holding pen | 50% |
| Fastening buttons | 77% |
| Opening jar | 50% |

The pen holding and button fastening tasks were included in the focus group discussion, and associated statements are summarised in Table 6. With both writing and buttoning, subthemes were related to problems in holding and strategies to get a good grip. A further subtheme emerged for buttoning. This was the issue of exerting sufficient pressure when buttoning up clothes.

**Table 6.** Focus group statements relating to hand activities - writing and doing up buttons.

| Group | ID | Time | Statement |
| --- | --- | --- | --- |
|  |  |  | **Writing** |
| A | 30 | 25:03 | It was quite tricky for me to concentrate on holding my pen properly and being able to write so I could understand what I'd actually written... prior to my chemo, I never had any trouble with my writing at all. |
| A | 25 | 28:88 | I found it difficult to write... and I thought that was a brain issue and a confidence issue... and I switched to a thicker pencil... perfect for me... It's got ridges in this section and I can feel the ridges... preferred implement for writing... because I can grip it better. |
| A | 23 | 31:16 | My hand tends to slip little bit more. I tend to grip that more so I think a thicker pen might be easier to write with... my grip is not as good as it used to be. |
|  |  |  | **Buttons** |
| A | 9 | 43:03 | Sometimes it can be really quite difficult to get the buttons, because it's a thicker material, ... it is two-handed, and it's ... getting it quite in the right place too, and getting the pressure. |
| A | 25 | 44:24 | What I do is to use kind of the pad of my finger rather than my finger tip to get a difficult button through a hole. I'm doing it now. I feel with the pad of my finger. I'm feeling the surface and the size of the button ... because I can't feel at the end. ... And then what I was doing was actually then tipping the button on its side and using my fingernail ... at one time during chemo and straight after, I couldn't really do buttons up. |
| A | 30 | 46:01 | I do find sometimes that it's quite difficult to do the button up on my shorts or my trousers... I don't recall having any trouble before, whereas I do now, particularly as I say, if my hands are cold. |
| A | 19 | 48:48 | it took a wee bit longer, and it was a bit more awkward when you were doing like buttons down the front of your shirt. ... But then I really did have problems with trying to do the cuffs, and it's a single handed action. ... and generally material is a bit thicker as well, and maybe the button hole is slightly tighter. |
| A | 23 | 49:29 | I noticed it more when my hands are cold ... So I do think there is a thing about blood flow or something. ... affect your pressure and doing the buttons and that type of thing. |
| B | 22 | 07:26 | I do use a buttonhole device to get dressed or alternatively buy clothing with stud or zip fasteners. |

Other manual activities affected by CIPN

Table 7 summarises the activities affected by CIPN (and associated percentages of respondents affected) captured by PNQ item 3 for the 14 participants who responded to this question. The results complement those for QLQ and highlight difficulty with buttons (plus other 2-handed manipulation activities involving buckles, shoelaces, jewellery and sewing). Difficulty in holding a pen for writing and using eating implements, all examples of single-handed tool use, also feature, but with lower proportions of respondents affected. Directed fingertip touch including using a touchscreen, phone, or keyboard constitutes another group of affected activities, again with lower proportions affected.

**Table 7.** Percentage of responses (N=14) to PNQ item 3 that indicated problems with manual activities involving bimanual manipulation (doing up buttons, buckles, lacing shoes, fastening jewellery, sewing), 1-hand tool use (writing, using eating implements), and directed touch (touchscreen, using phone, keyboard) activities. The corresponding numbers in each category, along with responses to non-dexterity items, can be found in S3 of the supplementary materials.

| **Class** | **Activity** |  |  |  |  |
| --- | --- | --- | --- | --- | --- |
| **Bimanual** | **Button** | **Buckle** | **Shoe laces** | **Jewellery** | **Sewing** |
| Percentage | 79% | 50% | 43% | 50% | 29% |
| **1-hand** | **Write** | **Fork** | **Knife** | **Spoon** |  |
| Percentage | 36% | 29% | 29% | 21% |  |
| **Directed** | **Touchscreen** | **Phone** | **Keyboard** |  |  |
| Percentage | 43% | 29% | 21% |  |  |

Insight into issues arising with manual activities comes from responses to UBQ item 5, “How does numbness/tingling affect activities?”. Table 8 shows impaired dexterity (in picking up and manipulating items) and dropping items (due to lower sensation) were reported by between half and a third of the participants. In these responses the participants often alluded to the loss of sensation, especially at the fingertips. Difficulty with achieving sufficient grip for both small (e.g. buttons) and large (e.g. jars) items was reported in 17% of cases. Some respondents attributed this to reduced finger strength. Several participants also reported slow response to temperature resulting in burns (13%) and painful sensitivity to cold (9%).

**Table 8.**
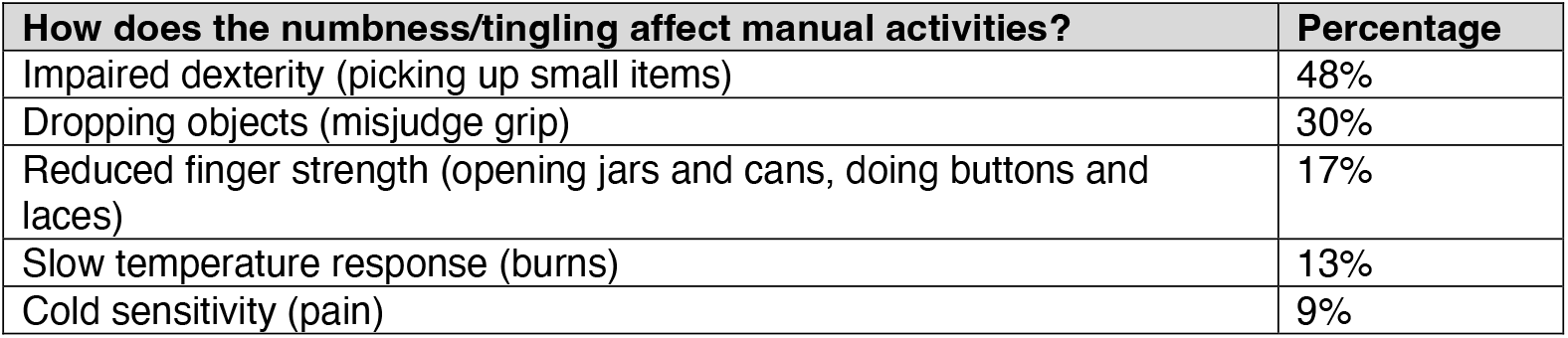
Percentage of respondents indicating issues in response to UBQ item 5: How does numbness/tingling affect activities?

A third of participants (N=8) provided responses to UBQ item 7, “What helps reduce feeling of numbness and tingling?” and UBQ item 8 “What strategies help reduce impact of numbness and tingling on your activities?”. In terms of reducing numbness, the suggestions they provided (as a proportion of those responding) were keeping hands warm (wearing gloves) (63%; N=5) and rubbing the skin (38%; N=3). With regard to strategies to help activities, two suggestions (25%) involved an aid (use a jar opener or button hook helper), two suggested using distraction or ignoring changes in sensation, and one each advised using muscle memory, moving slower, improving feel by using thicker wool (in knitting), and asking someone else to perform the task.

Various domestic tasks, ranging from washing dishes to knitting, were brought up in the Focus groups (see Table 9). Experiences included poor grip, especially with cold hands, shown under the theme of grip, and a feeling of clumsiness in fine manipulation, shown under the theme of dexterity, in Table 9.

**Table 9:** Focus group statements relating to grip failure and dexterity in other tasks involving the hands.

| Group | ID | Time | Statement |
| --- | --- | --- | --- |
|  |  |  | **Grip failure** |
| A | 25 | 38:08 | my husband said we needed to take out a bank loan for the amount of crockery that I dropped. Yes, I dropped a lot of things early on it doesn't happen now thankfully |
| A | 30 | 39:55 | When I come inside I have to wait for ages for my hands to warm up, ... to be able to be able to grab the shoe laces properly ... I can't do up my shoes if my hands are cold. Or I do them up and the laces come undone immediately. |
| A | 30 | 47:09 | if I'm just trying to open a plastic box ... for some reason, when my hands are cold, I just can't get enough pressure in them to open the box properly. I fumble with it. |
| B | 26 | 14:43 | I have dropped things before now because they (*fingers*) are numb. |
|  |  |  | **Dexterity** |
| A | 9 | 43:59 | (*when*) I'm reading the paper, ... separating pieces of paper much more difficult, even though I'm you know, I'm relatively recovered. |
| B | 11 | 02:03 | trying to do some fine knitting, and I was trying to count the stitches. I could hold the knitting needles, but I could not differentiate between the fine stitches with my fingertip (*participant 11 is blind*) ... changed to heavier wools to "see" what I was doing. |
| B | 10 | 10:28 | Initially, after the chemotherapy I did find my hands a little bit clumsy, but that quickly went away, and I have absolutely no problem with fine detail. I would say it was (over a period of) a few weeks, couple of months ... it was really just when I tried to do something small that required a bit of manipulation that I found my fingers a bit clumsy. |

## Discussion

Chemotherapy induced peripheral neuropathy (CIPN) is a common side-effect of chemotherapy with certain drugs, including taxanes and platinum analogues. The present study used patient reported outcome measures (PROM) questionnaires (QLQ, PNQ) with supplementary UBQ questions on the impact of CIPN on touch, on activities affected by impaired touch, and on strategies employed to reduce the impact of CIPN. In addition, focus groups explored pre-defined themes, including buttoning clothes, writing with a pen, and other manual tasks. The overall goal was to relate touch deficits, as well as the strategies used to cope with the deficits, to mechanisms of sensorimotor control. This was done partly to understand why some manual activities are more affected than others, but also to consider what further questions might be asked to help understand the problems people experiencing CIPN face with specific tasks. We also wanted to use the study findings to consider whether a test of sensorimotor function might further our understanding of the impact of CIPN on these manual activities.

The study included 25 participants treated on average 4.3 (SD=3.1) years previously with chemotherapy that induced CIPN. Twenty-three of the participants reported symptoms affecting the hands. This group were the focus of the current study. More than half of the participants reported that their symptoms of numbness and tingling were not changing or were getting worse. Persistent or worsening CIPN symptoms over similar durations, or longer, have previously been reported (Bao et al., 2016; Iveson et al., 2018; Winters-Stone et al., 2017). QLQ responses showed that, with similar proportions of participants reporting hand and foot effects of CIPN, more participants reported an impact on the performance of hand tasks. The tasks comprised doing up buttons, opening a jar and writing with a pen. The question then arises as to whether these tasks appear more sensitive to CIPN than the foot tasks because the manipulation elements of the hand tasks involve complex sensorimotor control where touch is more critical than in the foot tasks (which involved standing, walking and climbing stairs)? Alternatively, the hand and foot tasks might be equal in their dependence on touch, but the nature of the touch impairment in the hands has a greater impact. It would be interesting in future research to include questions that address other tasks in which foot touch sensation is more critical such as barefoot stepping in the shower or where the foot is involved in more manipulation-like activity, such as putting on socks. If this latter task where chosen it would be important to ask separately about the hand grip and foot aiming components.

From the perspective of sensorimotor control, it is interesting to ask what features the hand activities share and how they differ? The three activities where difficulty was reported in the QLQ questionnaire were holding a pen, fastening buttons and opening jars. These activities involve locating (initially with vision, then guided by tactile contact) and taking hold of an object (or material) between the tips or pads of the finger (or fingers) and opposed thumb. What contact forces are involved in taking hold, and how is tactile feedback used to regulate the forces? Sufficient grip force, directed into the object (buttonhole, pen, or jar top), is needed to keep stable contact and avoid the fingers slipping across (or even losing contact with) the object while applying force directed along the object surface. This latter tangential force causes the object to move against frictional resistance provided by a second separate object (button), or a connected object (jar) or surface (writing paper), held by the other hand (see Figures 1(b) and 2). Such frictional resistance is then registered as pressure and vibration on the contacting skin, providing information about the ongoing action and material properties of the object being handled.

Thus, each of the activities involves the generation of touch feedback, which can be used for grasping, and developing sufficient grip force to provide frictional resistance to the load forces involved in manipulation, which tend to cause the grasped object to slip. However, the manipulation forces have to be more accurately directed spatially in locating and inserting the button in the buttonhole and in shaping letters in writing compared to developing twist force (torque) between the all-digit grasp on the lid and power grip (palm and fingers together opposed by the thumb) on the jar. The contact in buttoning is more with the tips of the digits and involves both hands compared to finger pad manipulation with one hand in writing. Taking these points together, that is, the requirement for two hand manipulation, combined with the fact that CIPN involves the fingertips more than the finger pads, may be the reason that buttoning was found to be more frequently affected than writing. In the case of jar opening, the forces are higher than the other two tasks which may be a significant factor in the difficulty associated with CIPN, especially if it is combined with dry skin, which is commonly experienced after chemotherapy (Sibaud et al., 2016). Dry skin reduces friction and therefore increases the risk of the fingers slipping on the surface. This difficulty is compounded by CIPN effects on afferent fibres making it harder to detect slipping sensations which delays the corrective action.

The impact of CIPN on other manual activities was picked up by the PNQ. The activities identified fell into three groups. The first group, affecting higher proportions of participants, was bimanual manipulation (doing up buttons, buckles, lacing shoes, fastening jewellery, and sewing). The sensorimotor control challenges for these activities will be seen as overlapping those discussed above for the QLQ buttoning task. The last two activities in this group, involving jewellery and sewing, might be seen as being easier in that they require lower forces than the rest of the tasks in the group. However, this easing of difficulty is likely to be offset by the extra dexterity required with reduced tactile feedback due to the smallness of the items being gripped which, in turn, necessitates using the very tips of the digits where the loss of tactile sensation is more pronounced. The second and third PNQ activity groups were 1-hand tool use (writing, using eating implements), and directed touch (using touchscreen, phone, or keyboard), which involved lower proportions of participants. While sensorimotor control issues in 1-hand tool use may be seen as similar to those in writing, the nature of difficulties in the third PNQ group is less clear. One possibility is that they are related to difficulty in using touch to register when contact is made with response buttons (real or virtual), and when the response buttons have been depressed.

Further insight into the effects of CIPN on sensorimotor control of manual activities comes from the responses to UBQ. Impaired dexterity in picking up small items, as well as dropping items due to misjudged grip were reported. Both are consistent with reduced tactile feedback due to CIPN. Moreover, dropping crockery due to numbness was referred to as a specific example in the focus groups. Interestingly, difficulty with handling sheets of paper, for example turning pages, is noted as another specific example. Turning pages requires the tips of the fingers to gently rub and catch against the paper edge, which is likely to be difficult when, as in CIPN, tactile feedback from the fingertips is reduced.

One participant’s suggestion for coping with touch impairment in handling objects was to move slower. This might be expected to help by reducing acceleration, which reduces the likelihood of a slip, and also by giving more time for processing of tactile feedback to achieve corrective increases in grip and avoid dropping the object. In the case of writing, enlarged or ridged pen grip surfaces were reported to be helpful. This may be because the increased contact area and the ridges improved tactile feedback. In addition, both may have increased friction with the skin and so contributed greater resistance to slip. Other participant suggestions for reducing CIPN impact on manual activities included ignoring abnormal touch sensation and drawing more on muscle memory, both of which may change control from “closed” (with feedback) to “open” loop (no feedback) or, possibly, passes feedback control of the action from touch to vision.

Several participants reported problems with manual activities under the heading of general clumsiness. This suggests they were experiencing incoordination of finger movements relative to the object being manipulated. In future, it would be useful to explore whether such incoordination arises from muscle weakness or reduced tactile feedback. A similar issue was brought up in the focus groups, where reference was made to reduced finger strength in opening a box and to poor grip on shoelaces, in both cases with cold fingers. Again, it is not clear whether this was due to impaired tactile feedback or muscle weakness. It is also possible that chemotherapy side effects of dry skin (reducing friction between digits and object surfaces, so requiring higher grip force to prevent slipping), and fingernail damage (impairing manipulation at the very tips of the fingers) play a role. Another possibility is that there are deficiencies in proprioceptive input from muscle receptors, and this could also contribute to movement problems in CIPN. Deficits in proprioceptive guided control of movement including both upper and lower limbs, have been reported in CIPN (Wang et al., 2021). In future research, it will be important to investigate whether proprioceptive deficits may underlie reports of weakness, such as those noted in the present study.

The open-ended nature of the UBQ items generated useful hypotheses about the impact of CIPN on sensorimotor control of manipulation. In future research, it would be interesting to include questions focusing on specific problems identified in the present study. This might include asking with what implements and under what circumstances does dropping occur. However, questionnaire data, while capable of yielding valuable insights into participants’ experiences of CIPN, are prone to issues around subjective reporting, such as dependence on awareness, observation skills, recall abilities and response biases. One approach to these limitations is to use objective quantitative measures of tactile impairment to supplement PROMs. Molliasotis et al. (2019) used monofilaments to deliver controlled light touch pressure to determine detection thresholds as a measure of CIPN-related numbness. The incidence of patients exceeding criterion levels of numbness increased up to the end of chemotherapy and then declined in the following months. They reported parallel changes in the incidence of patients responding with above criterion levels of rated numbness on QLQ. Nielson et al. (2022) assessed vibration detection thresholds for hand and foot in addition to administering the QLQ. Average measures of both QLQ summed score and vibration thresholds showed a peak midway through chemotherapy treatment but also peaked again, 3-6 months after termination of therapy. In the latter study the authors also noted that the average performance curves for two different types of chemotherapy, taxanes and oxaliplatin, were similar.

Monofilament pressure and vibration detection threshold tests are important in reflecting the functioning of SA and RA afferents underlying tactile perception. The parallel nature of the PROMs and objective measures suggests that impairment in both pathways contribute to the experience of CIPN deficits reported by participants in the present study. Thus, it would make sense to include such tests in clinical research. However, our findings demonstrating the impact of sensory impairment on participants’ skilled actions suggest it would also be useful to include measures of action in order to assess the impact of CIPN on sensorimotor control. For example, a stylus-shaped object might be instrumented to assess grip and load forces when holding the stylus in precision grip and using the stylus tip to probe surface properties such as softness or texture. The same manipulandum might also be used to explore the ability to allow control slip of the object in the grasp, that is allowing the stylus to slide or rotate within a precision grip hold.

## Limitations

Although the present findings involved in-depth exploration of the participants’ experience, they were based on a relatively small number of self-selected participants and did not include age-matched controls. Additionally, CIPN was assessed at a single time point, asking participants to report both recent experiences (QLQ and PNQ) and those from years past (UBQ), with the latter constrained by recall biases. While it may be assumed that the findings will transfer to other similar contexts, such as people with similar clinical and demographic characteristics, further research is needed to examine, for example, generalisability to other contexts. Such work should expand on the present research, collecting data on CIPN experience over a wider time course, and gathering more detailed demographic data such as those regarding ethnicity, tumour type and treatment specifics and durations.

## Conclusion

Participants’ responses to questionnaires and themes developed in focus group have provided clear indications of the impact of touch impairment associated with CIPN on the performance of a range of everyday manual activities. Examination of the components of the affected activities, together with participants’ reports of alterations in their experience of touch, have provided hypotheses about why some tasks are more affected than other tasks.

An important element in coping with CIPN is an appreciation by patients and their clinicians of the nature of the disorder (Tanay et al., 2022). In this paper, we have worked towards a theoretically-based understanding of the sensorimotor issues that arise in CIPN. Sharing such understanding with those experiencing CIPN and their carers may be expected to contribute to an improved sense of involvement in the management of their symptoms. This, in turn, could help people with CIPN develop a sense of being empowered to explore strategies to overcome challenges posed by CIPN.

## Supporting information

Questionnaire responses and CIPN survey details

## Acknowledgements

The authors would like to extend their sincere thanks to the participants for generously sharing their time and insights, which have been invaluable in making this research possible.

## Data Availability

The data that support the findings of this study are available on request from the corresponding author (RDR).

