## Supplementary material for "Discriminative touch for manual action in chemotherapy induced peripheral neuropathy": Questionnaire responses and CIPN survey details

### Supplementary materials

These supplementary materials contain summaries of participant responses for each experience/grading of CIPN symptoms for (S1) the University of Birmingham Questionnaire (UBQ), (S2) the European Organization for Research and Treatment of Cancer Quality of Life (EORTC QLQ-CIPN20) Questionnaire (QLQ) and (S3) the Patient Neurotoxicity Questionnaires(PNQ). Details of the questionnaires presented in the online CIPN survey are shown in (S3). Questionnaire order reflects the proximity of the onset of the CIPN experience. UBQ covers the period from commencing chemotherapy to the present, QLQ the previous 7 days and PNQ the preceding 24 hours. Note, responses in S1-S3 include the data of two participants excluded from the main text analyses as their neuropathy was limited to the feet. Responses to open ended UBQ questions are available on request from the corresponding author.

**(S1) The number of participants selecting each multiple choice option for questions in the University of Birmingham Questionnaire (UBQ).**

| UBQ Questions | Number of Responses | | | |
|---|---|---|---|---|
| | **Yes** | **No** | | |
| **Item 2.** Did/do you have numbness and/or tingling as a result of chemotherapy? | 25 | 0 | - | - |
| | **Hands** | **Feet** | **Both** | |
| **Item 3a.** Does the numbness/tingling affect your Hands, Feet, Both, Other body parts (please specify below) | 6 | 2 | 17 | - |
| | **During chemotherapy** | | **After the end of chemotherapy** | |
| **Item 4.** When did the symptoms appear? | 18 | | 7 | |
| | **Handling objects** | **Balance** | **Both** | **Other** |
| **Item 5.** Does the numbness/tingling affect your | 12 | 1 | 9 | 3 |
| | **Getting worse** | **Getting better** | **Not changing** | |
| **Item 6.** Is the numbness/tingling.... | 3 | 6 | 16 | - |
| | **Yes** | **No** | | |
| **Item 7a.** Have you found any strategies or tricks that help reduce the impact of the numbness/ tingling on your activities? | 11 | 14 | - | - |
| | **Yes** | **No** | | |
| **Item 9a.** Did you, or do you have, any other symptoms related to chemotherapy on your hands/feet as well as the numbness/tingling (e.g. damaged fingernails)? | 11 | 14 | - | - |

**(S2) The number of participants selecting each option in the European Organization for Research and Treatment of Cancer Quality of Life Questionnaire (QLQ).**

| QLQ Questions | Number of Responses | | | |
|---|---|---|---|---|
| | Not at all (1) | A little (2) | Quite a bit (3) | Very much (4) |
| 1. Did you have tingling fingers or hands | 8 | 12 | 2 | 3 |
| 2. Did you have tingling toes or feet? | 10 | 6 | 4 | 5 |
| 3. Did you have numbness in your fingers or hands? | 9 | 10 | 2 | 3 |
| 4. Did you have numbness in your toes or feet? | 8 | 7 | 7 | 3 |
| 5. Did you have shooting or burning pain in your fingers or hands? | 18 | 3 | 1 | 3 |
| 6. Did you have shooting or burning pain in your toes or feet? | 17 | 6 | 0 | 2 |
| 7. Did you have cramps in your hands? | 18 | 4 | 1 | 2 |
| 8. Did you have cramps in your feet? | 14 | 9 | 1 | 1 |
| 9. Did you have problems standing or walking because of difficulty feeling the ground under your feet? | 20 | 3 | 1 | 1 |
| 10. Did you have difficulty distinguishing between hot and cold water? | 14 | 4 | 6 | 1 |
| 11. Did you have a problem holding a pen, which made writing difficult? | 13 | 10 | 1 | 1 |
| 12. Did you have difficulty manipulating small objects with your fingers (for example, fastening small buttons) | 7 | 12 | 2 | 4 |
| 13. Did you have difficulty opening a jar or bottle because of weakness in your hands? | 12 | 6 | 5 | 2 |
| 14. Did you have difficulty walking because your feet dropped downwards? | 24 | 0 | 0 | 1 |
| 15. Did you have difficulty climbing stairs or getting up out of a chair because of weakness in your legs? | 20 | 3 | 1 | 1 |
| 16. Were you dizzy when standing up from a sitting or lying position? | 16 | 8 | 0 | 1 |

| | | | | |
|---|---|---|---|---|
| 17. Did you have blurred vision? | 21 | 3 | 0 | 0 |
| 18. Did you have difficulty hearing? | 19 | 2 | 2 | 1 |
| Please answer the following question only if you drive a car<br><br>19.Did you have difficulty using the pedals? | 18 | 3 | 0 | 1 |
| Please answer the following question only if you are a man:<br><br>20. Did you have difficulty getting or maintaining an erection? | 3 | 3 | 0 | 1 |

**(S3) The number of participants selecting each option in the Patient Neurotoxicity Questionnaire (PNQ)**

| **Item 1.** | I have no numbness, pain or tingling in my hands or feet. | I have mild tingling, pain or numbness in my hands or feet. This does not interfere with my activities of daily living. | I have moderate tingling, pain or numbness in my hands or feet. This does not interfere with my activities of daily living. | * I have moderate to severe, tingling, pain or numbness in my hands or feet. This interferes with my activities of daily living. | * I have severe tingling, pain or numbness in my hands or feet. It completely prevents me from doing most activities of daily living. |
|---|---|---|---|---|---|
| **Number Selected** | 1 | 6 | 11 | 6 | 1 |

| **Item 2.** | I have no weakness in my arms or legs. | I have mild weakness in my arms or legs. This does not interfere with my activities of daily living. | I have moderate weakness in my arms or legs. This does not interfere with my activities of daily living. | * I have moderate to severe weakness in my arms or legs. This interferes with my activities of daily living. | * I have severe weakness in my arms or legs. It completely prevents me from doing most activities of daily living. |
|---|---|---|---|---|---|
| | 13 | 7 | 2 | 2 | 1 |

**Item 3.**

| If you have selected options marked with an asterisk (*) above, please indicate by selecting options or describing in the space provided below, which activity/activities have been interfered with as a result of chemotherapy. | | | |
|---|---|---|---|
| Button Clothes | **11** | Climb stairs | **3** |
| Use a knife | **4** | Type on a keyboard | **3** |
| Use a fork | **4** | Write | **5** |
| Use a spoon | **3** | Walk | **3** |
| Other eating utensils, etc | **1** | Put on jewellery | **7** |
| Open doors | **2** | Knit | **0** |
| Put in or remove contact lenses | **1** | Sew | **4** |
| Dial or use telephone | **4** | Work | **1** |
| Operation of remote control | **1** | Tie shoelaces | **6** |
| Fasten buckles | **7** | Drive | **2** |
| Sleep | **7** | Using touchscreens (e.g. smartphones, iPad/tablet) | **6** |

**(S4) CIPN Survey**

The survey is detailed below. It was presented online using MS Forms software and was in 4 sections: (A) Consent; name etc (B) UBQ questions (C) QLQ questions (D) PNQ questions (E) Contact details. Email contact was made subsequently with those who left email contact to ask for details of chemotherapy medication.

**Peripheral neuropathy and aging touch: Questionnaires on Coping with CIPN**

We would like to learn more about the experience of touch in people who have, or have had, chemotherapy-induced peripheral neuropathy (CIPN). There are 3 questionnaires in total to complete.

**A) University of Birmingham Consent**

Please complete the informed consent form below to indicate your agreement to participate before starting on the questionnaires.

1. Name:

2. Date of birth:

3. Handedness
   : Left
   Right
4. I have read the information sheet
   I have received enough information about the study
   I had a chance to ask questions
   I have received satisfactory answers to my questions
   I understand that I am free to leave the study anytime without having to give a reason

   Signature:

   Date:

#### B) University of Birmingham Questionnaire (UBQ) on symptoms of CIPN

This first questionnaire below is concerned with understanding if you have ever had symptoms of CIPN, how they have affected touch in your hands and/or feet, what impact this has had on your activities, and what helps to reduce the impact.

1. When did you finish your chemotherapy?

2. Did/do you have numbness and/or tingling as a result of chemotherapy?

   If No please skip to the next questionnaire by scrolling to the end of this page and clicking Next.
   Yes No

3a. Does the numbness/tingling affect your
Hands
Feet
Both
Other body parts (please specify below)

3b. How much of your body part is affected by the numbness/tingling?

4. When did the symptoms appear?
During chemotherapy
After the end of chemotherapy

5a. Does the numbness/tingling affect your
Handling objects
Balance
Both
Other

5b. How does the numbness/tingling affect these activities?

6. Is the numbness/tingling....
Getting worse
Getting better
Not changing

7a. Have you found anything that helps reduce the feeling of numbness/tingling?

7b. Please describe what helps.

8a. Have you found any strategies or tricks that help reduce the impact of the numbness/tingling on your activities?
Yes No

8b. Please describe these strategies.

9a. Did you, or do you have, any other symptoms related to chemotherapy on your hands/feet as well as the numbness/tingling (e.g. damaged fingernails)?
Yes No

9b. Please describe the other symptoms.

**D) European Organization for Research and Treatment of Cancer Quality of Life Questionnaire (EORTC QLQ-CIPN20)**

Patients sometimes report that they have the following symptoms or problems. For this third questionnaire, please indicate the extent to which you have experienced these symptoms or problems during the past week. Please answer by selecting the option that best applies to you.

During the past week:

| | Not at all | A little | Quite a bit | Very much |
|---|---|---|---|---|
| 1. Did you have tingling fingers or hands | 1 | 2 | 3 | 4 |
| 2. Did you have tingling toes or feet? | 1 | 2 | 3 | 4 |
| 3. Did you have numbness in your fingers or hands? | 1 | 2 | 3 | 4 |
| 4. Did you have numbness in your toes or feet? | 1 | 2 | 3 | 4 |
| 5. Did you have shooting or burning pain in your fingers or hands? | 1 | 2 | 3 | 4 |
| 6. Did you have shooting or burning pain in your toes or feet? | 1 | 2 | 3 | 4 |
| 7. Did you have cramps in your hands? | 1 | 2 | 3 | 4 |
| 8. Did you have cramps in your feet? | 1 | 2 | 3 | 4 |
| 9. Did you have problems standing or walking because of difficulty feeling the ground under your feet? | 1 | 2 | 3 | 4 |
| 10. Did you have difficulty distinguishing between hot and cold water? | 1 | 2 | 3 | 4 |
| 11. Did you have a problem holding a pen, which made writing difficult? | 1 | 2 | 3 | 4 |
| 12. Did you have difficulty manipulating small objects with your fingers (for example, fastening small buttons) | 1 | 2 | 3 | 4 |
| 13. Did you have difficulty opening a jar or bottle because of weakness in your hands? | 1 | 2 | 3 | 4 |

| | | | | |
|---|---|---|---|---|
| 14. Did you have difficulty walking because your feet dropped downwards? | 1 | 2 | 3 | 4 |
| 15. Did you have difficulty climbing stairs or getting up out of a chair because of weakness in your legs? | 1 | 2 | 3 | 4 |
| 16. Were you dizzy when standing up from a sitting or lying position? | 1 | 2 | 3 | 4 |
| 17. Did you have blurred vision? | 1 | 2 | 3 | 4 |
| 18. Did you have difficulty hearing? | 1 | 2 | 3 | 4 |
| Please answer the following question only if you drive a car<br>19.Did you have difficulty using the pedals? | 1 | 2 | 3 | 4 |
| Please answer the following question only if you are a man:<br>20. Did you have difficulty getting or maintaining an erection? | 1 | 2 | 3 | 4 |

**C) Patient Neurotoxicity Questionnaire (PNQ)**
For this second questionnaire, please select the option which best describes your symptoms today.

**Item 1.**

| I have no numbness, pain or tingling in my hands or feet. | I have mild tingling, pain or numbness in my hands or feet. This does not interfere with my activities of daily living. | I have moderate tingling, pain or numbness in my hands or feet. This does not interfere with my activities of daily living. | * I have moderate to severe, tingling, pain or numbness in my hands or feet. This interferes with my activities of daily living. | * I have severe tingling, pain or numbness in my hands or feet. It completely prevents me from doing most activities of daily living. |
|---|---|---|---|---|

**Item 2.**

| I have no weakness in my arms or legs. | I have mild weakness in my arms or legs. This does not interfere with my activities of daily living. | I have moderate weakness in my arms or legs. This does not interfere with my activities of daily living. | * I have moderate to severe weakness in my arms or legs. This interferes with my activities of daily living. | * I have severe weakness in my arms or legs. It completely prevents me from doing most activities of daily living. |
|---|---|---|---|---|

**Item 3.**

| If you have selected options marked with an asterisk (*) above, please indicate by selecting options or describing in the space provided below, which activity/activities have been interfered with as a result of chemotherapy. | | |
|---|---|---|
| Button Clothes | | Climb stairs |
| Use a knife | | Type on a keyboard |
| Use a fork | | Write |
| Use a spoon | | Walk |
| Other eating utensils, etc | | Put on jewellery |
| Open doors | | Knit |
| Put in or remove contact lenses | | Sew |
| Dial or use telephone | | Work |
| Operation of remote control | | Tie shoelaces |
| Fasten buckles | | Drive |

| | | |
|---|---|---|
| Sleep | | Using touchscreens (e.g. smartphones, iPad/tablet) |

**E) Thank you for your time.**

If you would like to take part in an on-line focus group to follow up on these questions or to participate in touch sensory testing in the lab, please indicate below and we will get in touch with you.

Please contact me about taking part in:
An on-line touch focus group
Touch sensory testing in the lab
Both

I am providing my email and/or phone number for this purpose.

My name is
